# DICE: Deep Significance Clustering for Outcome-Driven Stratification

**DOI:** 10.1101/2020.10.04.20204321

**Authors:** Yufang Huang, Joel C. Park, Kelly M. Axsom, Lakshminarayanan Subramanian, Yiye Zhang

## Abstract

We present deep significance clustering (DICE), a framework for jointly performing representation learning and clustering for “outcome-driven” stratification. Motivated by practical needs in medicine to risk-stratify patients into subgroups, DICE brings self-supervision to unsupervised tasks to generate cluster membership that may be used to categorize unseen patients by risk levels. DICE is driven by a combined objective function and constraint which require a statistically significant association between the outcome and cluster membership of learned representations. DICE also performs a neural architecture search to optimize cluster membership and hyper-parameters for model likelihood and classification accuracy. The performance of DICE was evaluated using two datasets with different outcome ratios extracted from real-world electronic health records of patients who were treated for coronavirus disease 2019 and heart failure. Outcomes are defined as in-hospital mortality (15.9%) and discharge home (36.8%), respectively. Results show that DICE has superior performance as measured by the difference in outcome distribution across clusters, Silhouette score, Calinski-Harabasz index, and Davies-Bouldin index for clustering, and Area under the ROC Curve for outcome classification compared to baseline approaches.

## 1 Introduction

Representation learning [1, 2] and clustering [3] are unsupervised algorithms whose results are driven by input features and priors. They are often exploratory in nature, but in certain use cases users have *a priori* expectations for the outputs from representation learning and clustering. In the latter case, having targeted self-supervision in the learning process so as to meet the expectation of the users brings practical value for representation learning and clustering algorithms. This paper proposes deep significance clustering (DICE), an algorithm for self-supervised, interpretable representation learning and clustering that targets features that best stratify a population concerning specific outcomes of interest.

We are motivated by a practical need in medicine to identify and understand characteristics of subgroups of patients [4, 5, 6]. Patient health information is extremely complex, capturing high-dimensional, temporal, and heterogeneous data on diseases, biomarkers, medications, procedures, among other health indicators. For instance, heart failure (HF) is a syndrome that impacts nearly 6 million Americans and is associated with a 50% 5-year mortality [7]. More than 80% of individuals suffer from three or more comorbidities [8]. There are proven HF therapies that prolong survival. For patients in a moderately sick cohort, known as Stage C, therapies are oral medications, known as neurohormonal blockade. This cohort of patients have a 5-year survival of 75%. For the sickest cohort of heart failure patients, known as Stage D or end stage HF, therapies are focused on heart replacement like heart transplant or left ventricular assist devices. This population has a 20% 5-year survival [7]. On the contrary, therapeutic guidelines for novel coronavirus disease 2019 (COVID-19) are not as clearly delineated as HF treatment. In both scenarios, the complexity due to frequent comorbidity or the lack of clear guidelines warrant the discovery of patient subtypes to assist with clinical decision making. While clustering is an obvious choice for patient subtyping, the heterogeneous health data present a challenge to such unsupervised algorithm for its incapability to elicit cluster membership that is targeted for an outcome of interest, such as whether a HF patient can be safely discharged home and the mortality risk of COVID-19 patients. This challenge arises because a regular clustering algorithm may find clusters of patients who differ with respect to factors that are not related to meaningful clinical endpoints.

Addressing such clinical needs, DICE, a framework to learn a deep representation and cluster memberships from heterogeneous data was developed in an effort to bridge representation learning, clustering, and targeted outcome separation. Its architecture is illustrated in Fig. 1. Representation learning allows us to discover a concise representation from the heterogeneous and sparse health data, which we use to discover latent clusters within a patient population with clustering algorithms. As a way to provide more interpretability to the representation learning and clustering, DICE uses a combined objective function and a constraint that requires statistically different outcome distribution across clusters. The statistical significance is determined using models that are well-understood by clinicians such as regression while adjusting for patient demographics. The combined objective function and constraint serve to force DICE to learn representations that lead to clusters that are discriminative to the outcome of interest. Furthermore, a neural architecture search (NAS) is designed with an alternative grid search for the number of clusters and hyper-parameters in the representation learning. The finalized representation and cluster memberships, which represent significantly different outcome levels, are then used as the class labels for a multi-class classification. This is intended to allow new patients to be categorized according to risk-level specific subgroups learned from historic data.

**Figure 1:**
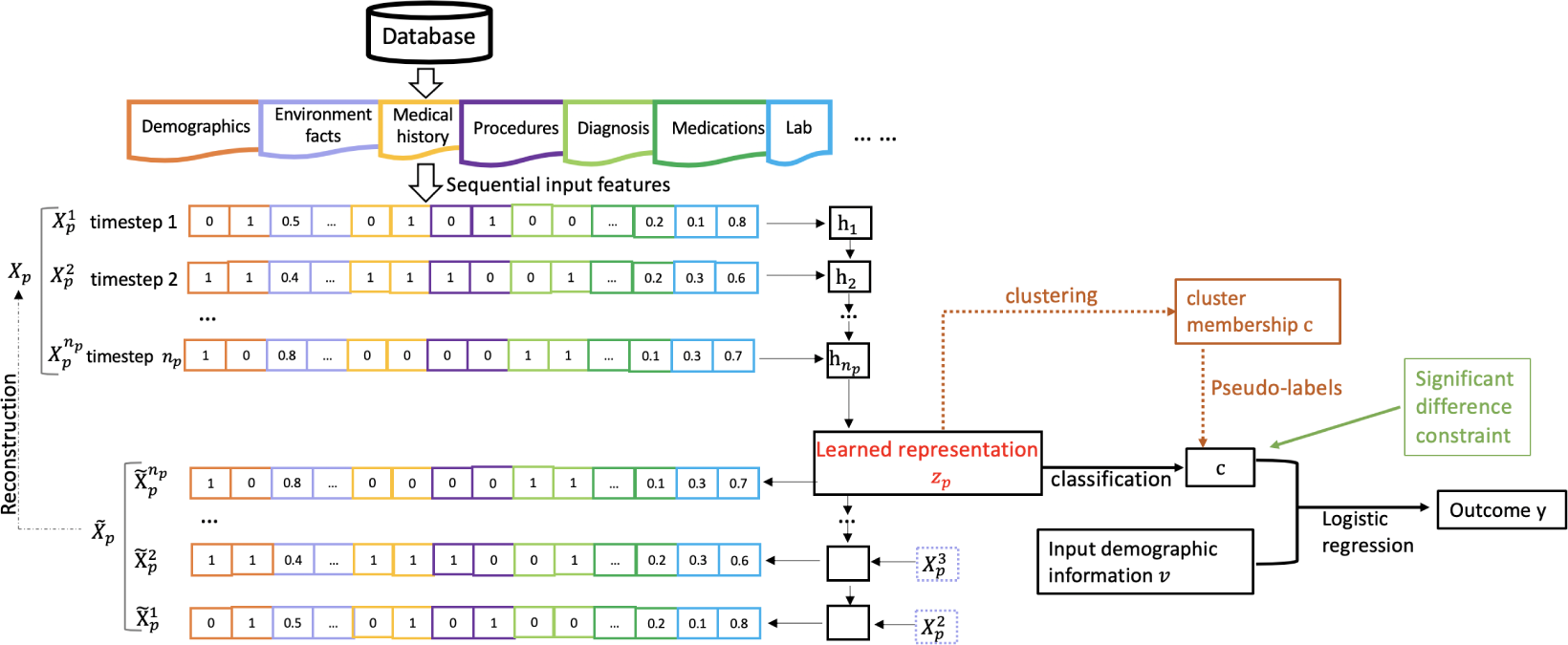
The framework of the proposed deep significance clustering (DICE). Clustering is applied to the representation **z**_*p*_. A statistical significance constraint is explicitly added to ensure the association of the clustering membership **c** and outcome *y* to facilitate the learning of discriminative representations **z**_*p*_.

Previous studies [9] that incorporated statistical significance analyzed it separately after the representation learning process. Our paper considers the statistical significance while performing deep clustering as a constraint. To summarize, our approach makes the following key contributions:

- We propose a combined objective function to achieve the joint optimization for outcome-driven representation and clustering membership from heterogeneous health data.
- We propose an explicit constraint that forces statistical significance of the association between the cluster membership with the outcome to drive learning.

We evaluated DICE on two real-world datasets collected from electronic health records (EHR) data at an academic medical center. Extensive experiments and analyses demonstrate that the DICE obtains better performance than several baseline approaches in outcome discrimination, Area under ROC Curve (AUC) for prediction, and clustering performance metrics including Silhouette score, Calinski-Harabasz index and Davies-Bouldin index.

## 2 Related Work

Clustering is a fundamental topic in the exploratory data mining which can be applied to many fields, including bioinformatics [10], marketing [11], computer vision [12] and natural language processing [13]. Due to the inefficiency of similarity measures with high-dimensional big data, traditional clustering approaches, e.g., *k*-means [14], finite mixture model [15, 16] and Gaussian Mixture Models (GMM) [17], generally suffer from high computational complexity on large-scale datasets [18]. Also, the mixture models have distribution assumptions on observations [19]. Jagabathula [20] proposed a conditional gradient approach for nonparametric estimation of mixing distributions. Data transform approaches which map the raw data into a new feature space have been studied, including principal component analysis (PCA) [21], kernel methods [22], model-based clustering [23, 19] and spectral methods [24, 25]. However, clustering of high-dimensional heterogeneous data is still challenging for these approaches because of inefficient data representation. Deep representation learning can be used to transform the data into clustering-friendly representation [26, 27, 28]. Parametric t-SNE [29] uses deep neural network to parametrize the embedding of t-SNE [30] with the same time complexity of *O*(*n*^2^), where *n* is the number of data points. DEC [27] further relaxes parametric t-SNE with a centroid-based probability distribution which reduces complexity to *O*(*nk*) from tree-based t-SNE of *O*(*n*log(*n*)), where *k* is the number of centroids. Some approaches learn self-supervised representation [31, 32, 33]. Recent deep clustering approaches are learning-based and conduct inference in one shot, consisting of two stages, i.e., deep representation learning followed by various clustering models. Caron et al. [33] jointly learned the parameters of a deep network and the cluster assignments of the resulting representation. DGG [12] further uses gaussian mixture variational autoencoders and graph embedding to improve the clustering and data representation abilities. DICE considers statistical significance and proposes a novel constraint to obtain statistical significant clustering results.

## 3 Method

Given a dataset 𝕏 = {**X**_1_, …, **X**_*P*_} with *P* subjects, we denote each subject as a sequence of events 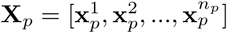 of length *n*_*p*_. A multivariate feature vector 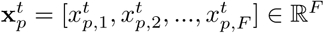 is the *t*-th instance of subject *p* in sequence **X**_*p*_, where *F* is the number of features at each timestamp. We have an outcome *y*_*p*_ for each subject *p*. Our goal is to stratify 𝕏 of *P* subjects into *K* clusters while enforcing statistical significance in the association of the cluster membership and the outcome while adjusting for relevant covariates.

### 3.1 Learning representation

The first step is to transform discrete sequences into latent continuous representations, followed by clustering and classification. The latent representation learning for each subject is performed by an LSTM autoencoder (AE) [34, 35, 36]. The AE consists of two parts, the encoder and the decoder, denoted as ℰ and ℱ, respectively. Given the *p*-th input sequence 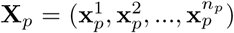, the encoder can be formulated as **z**_*p*_ = ℰ (**X**_*p*_; *θ*_ℰ_), where **z**_*p*_ ∈ ℝ^*d*^ is the representation, and ℰ is a LSTM network with parameter *θ*_ℰ_ [37]. We choose the last hidden state **z**_*p*_ of LSTM to be the representation of the input **X**_*p*_. The decoder can be formulated as 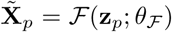, and ℱ is the other LSTM network with parameter *θ*_ℱ_. The representation learning is achieved by minimizing the reconstruction error

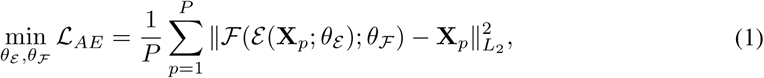

where we use *L*_2_ norm in the loss.

### 3.2 Self-supervised learning by clustering

The obtained representations 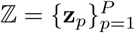 can be employed for clustering with *K* clusters,

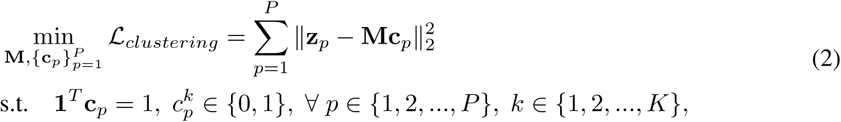

where *K* is a hyper-parameter to tune, 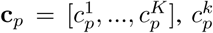 is cluster membership of cluster *k*, **M** ∈ ℝ^*d×K*^ and the *k*-th columns of **M** is the centroid of the *k*-th cluster.

To enable fast inference, we build a learning-based deep clustering based on self-supervision from **c**_*p*_ in equation (2). Please note that we can utilize other priors of equation (2) in the DICE. We employ the clustering results 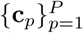 from *a priori* in equation (2) as pseudo-labels, and update the parameters of the encoder ℰ and ℱ. The cluster membership assignment can be formulated as a classification network.

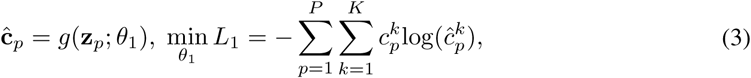

where 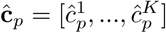 is the predicted cluster membership from the classification network *g*(·; *θ*_1_), *θ*_1_ is the parameter in the classification network, *L*_1_ is the negative log-likelihood loss for multi-class classification.

The deep clustering bridges the representation learning with the following statistical significance constraint related to the outcome.

For inference, we assign the cluster membership through equation (2) with fixed **M** from training.

### 3.3 Statistical significance constraint

We propose a novel statistical significance constraint to the clustering membership w.r.t. the outcome distribution while adjusting for relevant covariates in the DICE. After obtaining cluster memberships 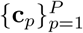 for *K* clusters, we impose a statistical significance constraint on the cluster membership to drive the representation learning. We fuse the statistical significance constraint into our neural network.

First, in our neural network, we use the cluster memberships and non-clinical events like demographic features to predict outcome, formulated as:

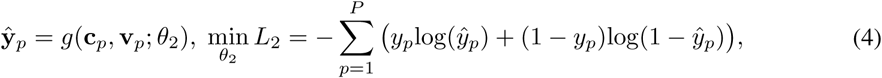

where **v**_*p*_ represents non-clinical events, *g*(·; *θ*_2_) is the logistic regression, *L*_2_ is the negative log-likelihood loss for binary classification problem.

Second, to quantify the significant difference of cluster *k*_1_ and cluster *k*_2_ (*k*_1_ ≠ *k*_2_), we use likelihood-ratio test [38] to calculate the *p*-value of variable 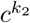 when considering cluster 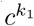 as the baseline, where *c*^*k*^ refers to the cluster membership belonging to cluster *k*. We denote the likelihood-ratio as 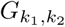, then obtain the *p*-value from Chi-square distribution, denoted as 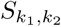. Finally, we have a matrix **S** ∈ ℝ^*K×K*^ with 0 as diagonal elements, and *S*_*k*1,*k*2_(*k*_1_ ≠ *k*_2_) is the *p*-value represent the significance difference of cluster *k*_2_ corresponding to baseline cluster *k*_1_. If all the elements in **S** are below a predefined threshold of significance *α* (equivalently, 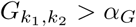), we conclude that all the clusters are significantly different with each other related to outcome *y*.

For implementation, we use mask technique to mask each variable of input **c**_*p*_, then calculate the likelihood ratio 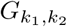 and add significance constraint on 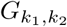, that is 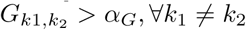.

### 3.4 Objective function

We use NAS approaches to obtain our final model. The first is weight optimization of a given network architecture, which in our method is the network architecture with fixed clusters number *K* and hidden state dimension *d*. The second is the neural architecture search process.

#### 3.4.1 Optimization of a given network architecture

We denote our network architecture as 𝒩 (*K, d, θ*), where {*θ*_ℰ_ = *θ*_ℱ_, *θ*, **M**, *θ*_1_, *θ*_2_} are the weights of network. The optimization problems is

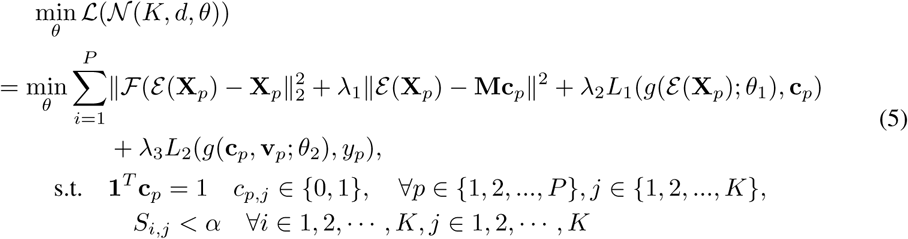

To implement, we iteratively optimize the clustering and the other components with the statistical significance constraint. That is,

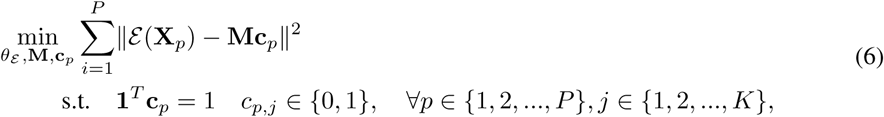

and

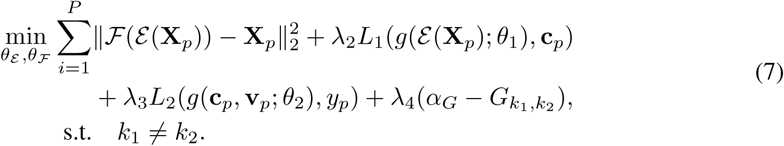

The algorithm is elaborated in Algorithm 1.

##### Algorithm 1: Deep significance clustering

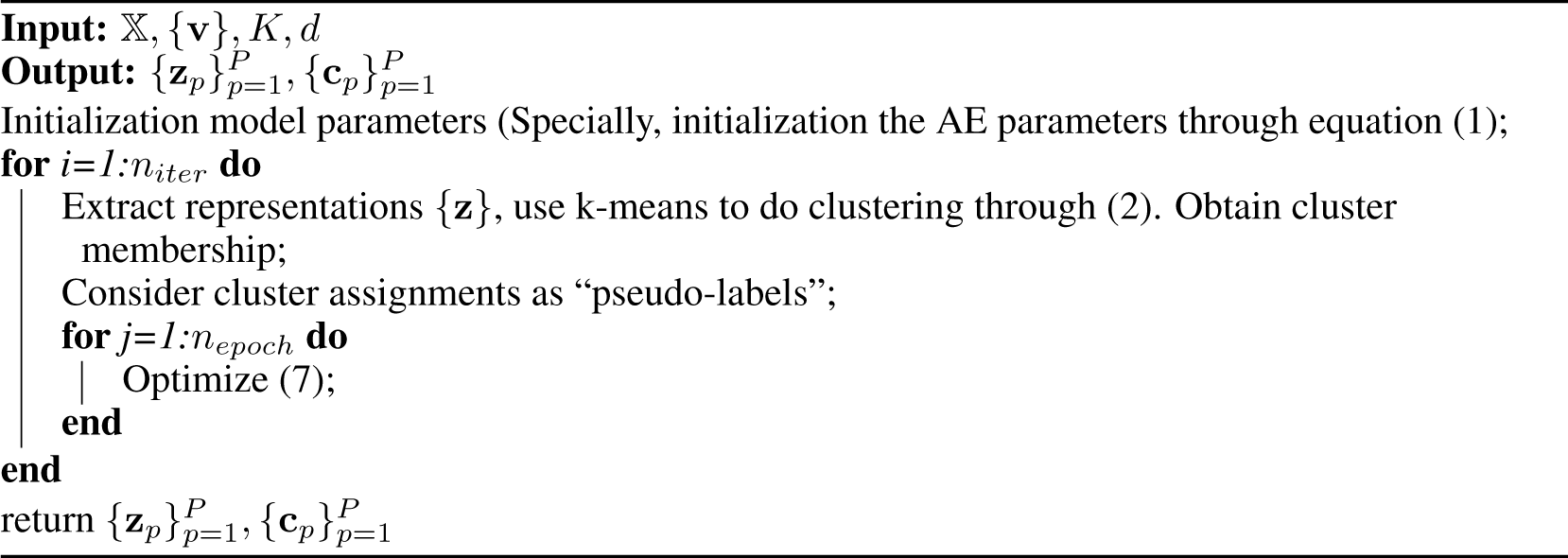

#### 3.4.2 Architecture search

We choose the architecture which is trained on the training set and has the best evaluation performance on validation set, that is

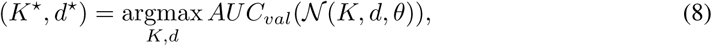

where *AUC*_*val*_(·) is the AUC score on the validation set.

## 4 Experiments

We conducted experiments on two datasets and compared against 3 baseline methods. We also carried out ablation experiments to study the impact of statistical significance constraint of DICE.

### 4.1 Experimental setting

#### Data

We used datasets on two patient populations: HF and COVID-19, extracted from electronic health records (EHRs) at an urban academic medical center. The datasets were split into training, validation, and test sets in a 4 : 1 : 1 ratio.

- **HF**: We included HF patients (*n* = 1585) aged 18 to 89 from years 2014 to 2018 who were treated on the Medicine service. HF was defined by ICD-9/10-CM. Only patients whose initial and final diagnoses both contained HF were included to ensure that HF was treated during the hospital stay. The outcome is defined as discharged to home (36.8%). Sequential medical orders were included in the data.
- **COVID-19**: We included patients aged 18 to 101 who were presented to the emergency department and admitted for COVID-19 (*n* = 968) in 2020. COVID-19 was defined by a positive polymerase chain reaction test. The outcome is in-hospital mortality (15.9%). Age, race, and sequential laboratory values were included in the data.

#### Baselines

We compared our method with baseline method including (1) principal component analysis (PCA) [21], (2) AE, and (3) AE + classification. In (2) AE, clustering was applied directly to representations learned from AE [36]. In (3) AE + classification, first we jointly trained AE and classification with representation learned from AE as the input for classification, then applied clustering to the final learned representation. No statistical constraint or NAS was used in training the baselines, but we report below the results with the same dimension of representation and cluster number with DICE. For AE, we chose the minimum reconstruction error on validation set. For AE + classification, we chose the results with the maximum AUC on the validation set.

#### Training

We conducted experiments in PyTorch framework on NVIDIA GeForce RTX 2070. We initialized the model parameters of AE by 1 epoch training. We set *α* = 0.05, *α*_*G*_ = 3.841, *n*_*iter*_ = 60, *n*_*epoch*_ = 1. The *λ*_1_, *λ*_2_, *λ*_3_ were set as 0.1, 10, 1.0 respectively based on the performance on the validation set. It took approximately 7 minutes for each result with fixed neural architecture.

### 4.2 Results

We used NAS to choose the best model, then qualitatively assessed our method with baselines using clustering and classification metrics. Ablation studies were also conducted to compare performance absent the statistical significance constraint.

#### Neural network architecture search

Our search spaces were {(*K, d*)|*K* ∈ {2, 3, 4, 5}, *d* ∈ {20, 25, …, 100}} for the HF dataset and {(*K, d*)| *K* ∈ {2, 3, 4, 5}, *d* ∈ {10, 11, …, 30}} for the COVID-19 dataset, which are set according to the number of features and size of datasets. Figure 2 demonstrates the NAS process, with AUC values from the validation set of different neural network architecture on the Y-axis and *d* on the X-axis. From Figure 2, we can see that the statistical significance constraint can drive the model towards higher AUC, as also demonstrated in the ablation study described below. Maximizing the model AUC, *K* = 4, *d* = 35 for the HF dataset, and *K* = 3, *d* = 21 for the COVID-19 dataset, were chosen as the optimal parameters.

**Figure 2:**
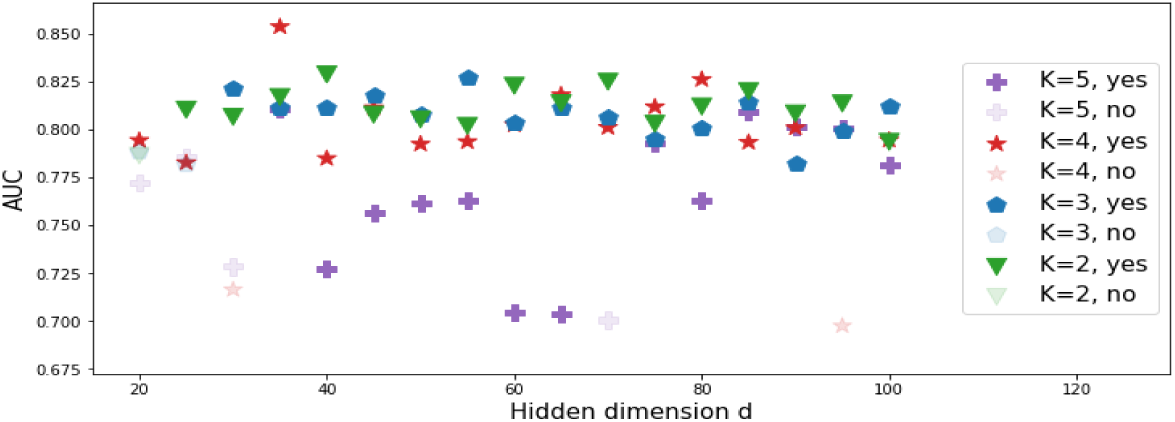
The model selection on HF dataset. “yes” represents that the architecture network met the significance constraint, and “no” otherwise.

#### Visualization of representation

For the HF dataset, we demonstrate the clustering results through the visualization of representation in Figure 3. Compared with Figure 3(b), Figure 3(c) and Figure 3(d), the 4 clusters in Figure 3(a) discovered by DICE displayed tighter separation, with the highest outcome ratio 79.93% in cluster 1 to the lowest outcome ratio 8.61% in cluster 4. The baseline AE+classification also discovered 4 clusters with the outcome ratio in each cluster ranging from 72.22% to 5.85%, but the clusters are not well separated. PCA and AE did not discover clusters with outcomes as clearly separated as DICE, likely because the baseline from those two baselines are not outcome-driven. Our DICE learns representation through outcome-driven and self-supervised learning from pseudo-labels, therefore we can obtain clear outcome risk stratification and well separated clusters at the same time. Results for the COVID-19 dataset are shown in Figure 4. DICE again obtained clearer separation between clusters and outcome stratification as measured by the difference in outcome ratio within each cluster.

**Figure 3:**
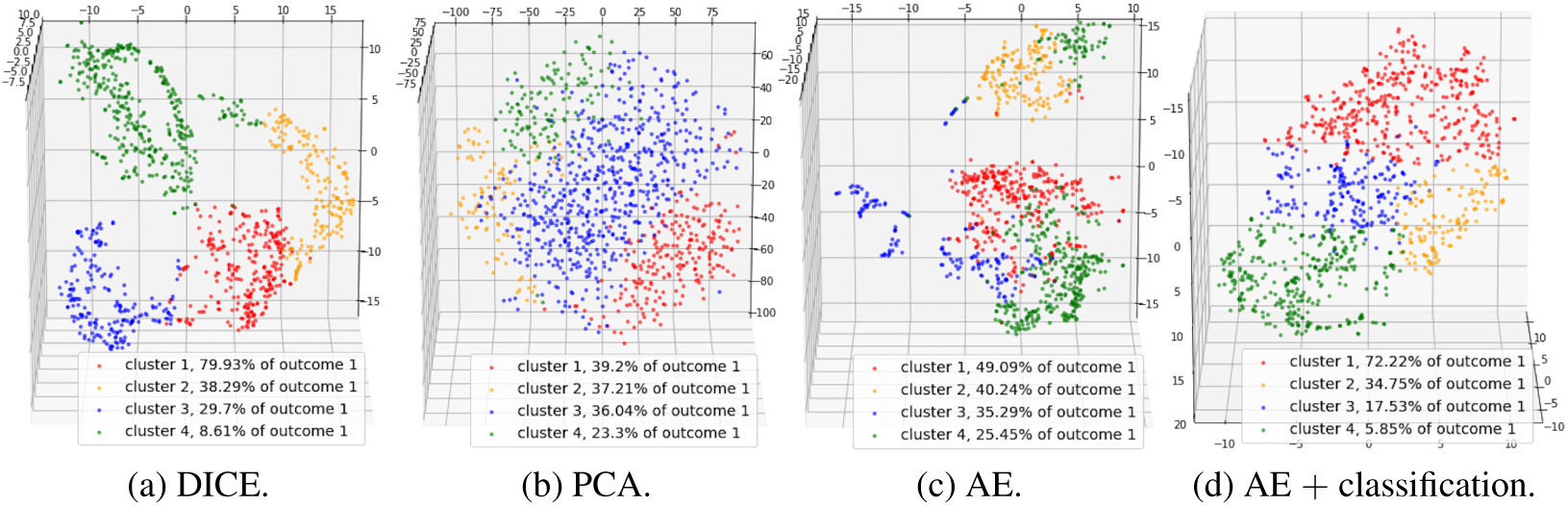
Visualization of patient subtyping results by various methods on HF dataset.

**Figure 4:**
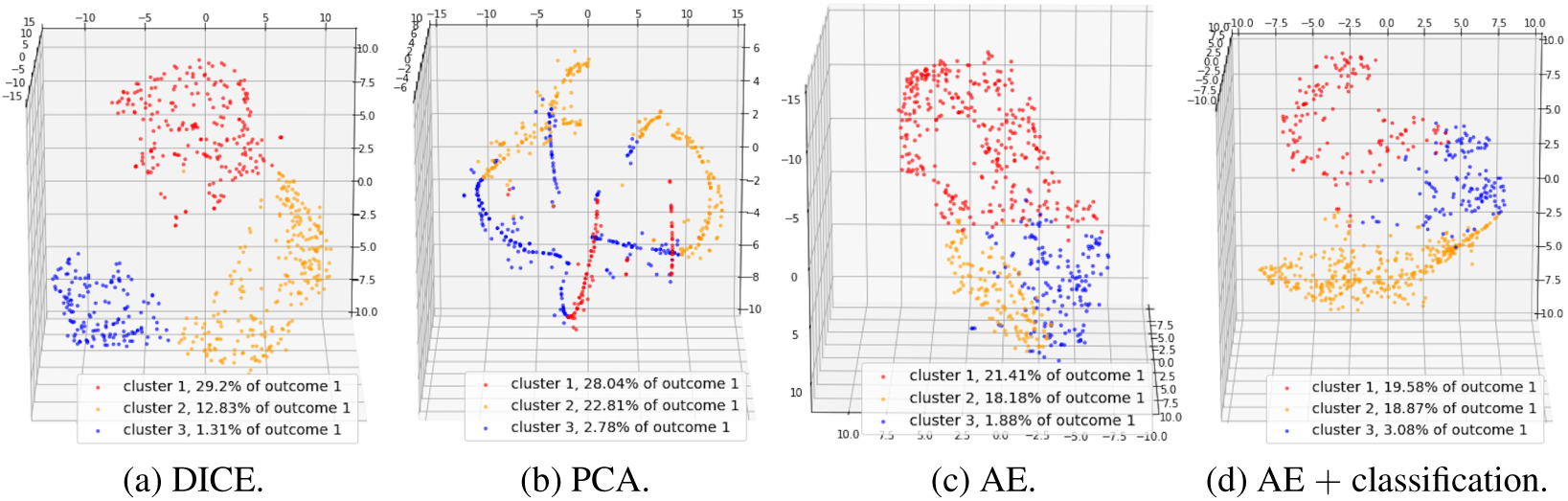
Visualization of patient subtyping results by various methods on COVID-19 dataset.

#### Clustering performance on unseen data

The learned cluster membership from historic data can serve as a pseudo-label for unseen data, such that new patients may be classified into one of the risk levels. The clustering performance on the test set is shown in Table 1. Since the ground truth labels of stratification are unknown, we used Silhouette score [39], Calinski-Harabasz index [40], and Davies-Bouldin index [41] to evaluate the clustering performance. DICE achieved the best separation across the three metrics in HF dataset, and outperforms two out of three metrics in the COVID-19 dataset. DICE underperformed to AE + classification in Calinski-Harabasz index in the COVID-19 dataset. Compared to HF, the COVID-19 population has a much diverse health conditions, which may have presented a challenge to minimize within-cluster variance.

**Table 1:**
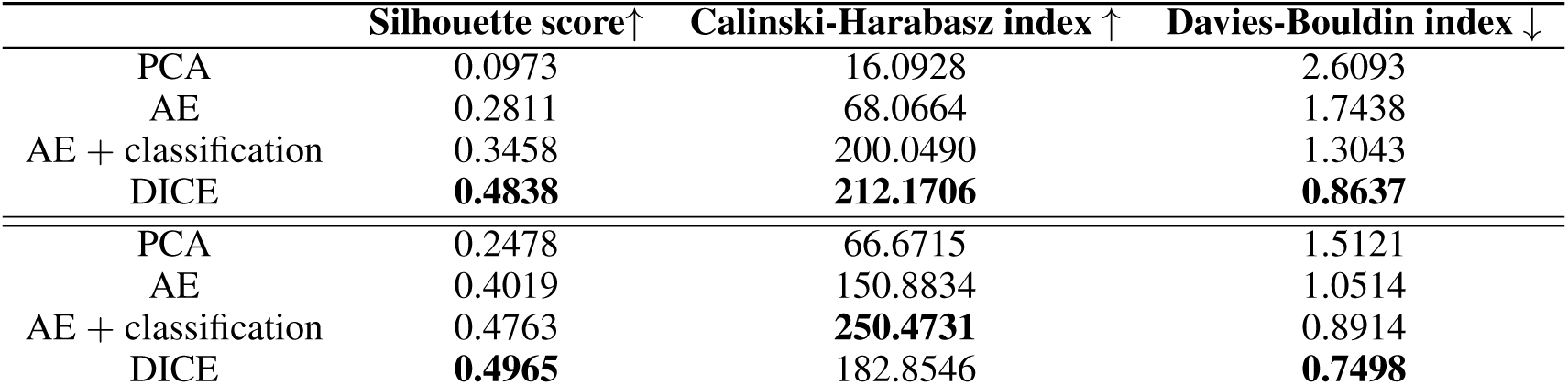
Clustering performance evaluation on test set. Upper: HF dataset. Lower: Covid-19 dataset.

| | Silhouette score↑ | Calinski-Harabasz index ↑ | Davies-Bouldin index ↓ |
| --- | --- | --- | --- |
| PCA | 0.0973 | 16.0928 | 2.6093 |
| AE | 0.2811 | 68.0664 | 1.7438 |
| AE + classification | 0.3458 | 200.0490 | 1.3043 |
| DICE | **0.4838** | **212.1706** | **0.8637** |
| PCA | 0.2478 | 66.6715 | 1.5121 |
| AE | 0.4019 | 150.8834 | 1.0514 |
| AE + classification | 0.4763 | **250.4731** | 0.8914 |
| DICE | **0.4965** | 182.8546 | **0.7498** |

#### Outcome classification via learned representation

We used the learned representation from DICE for outcome classification using logistic regression, as shown in Table 2. DICE outperformed the baselines in AUC, accuracy, true positive rate, false negative rate, and negative predictive value. The reason DICE had high FPR and low TNR and PPV compared to baselines may be explained by the high negative case ratio in both datasets.

**Table 2:** Outcome prediction comparison. Upper: HF dataset, Lower: COVID-19 dataset.

| | AUC↑ | ACC↑ | FPR↓ | TPR↑ | FNR↓ | TNR↑ | PPV↑ | NPV↑ |
| --- | --- | --- | --- | --- | --- | --- | --- | --- |
| PCA | 0.773 | 0.712 | 0.222 | 0.598 | 0.402 | 0.778 | 0.611 | 0.769 |
| AE | 0.712 | 0.697 | **0.150** | 0.433 | 0.567 | **0.850** | 0.627 | 0.721 |
| AE + classification | 0.818 | 0.765 | 0.251 | 0.794 | 0.206 | 0.746 | 0.647 | 0.862 |
| DICE | **0.834** | **0.780** | 0.257 | **0.845** | **0.155** | 0.743 | **0.656** | **0.892** |
| PCA | 0.757 | 0.772 | 0.221 | 0.731 | 0.269 | 0.779 | 0.387 | 0.938 |
| AE | 0.855 | 0.840 | 0.147 | 0.769 | 0.231 | 0.853 | 0.5 | 0.951 |
| AE + classification | 0.889 | 0.877 | **0.051** | 0.5 | 0.5 | **0.949** | **0.65** | 0.908 |
| DICE | **0.907** | **0.889** | 0.096 | **0.808** | **0.192** | 0.904 | 0.618 | **0.961** |

#### Fairness on race

To ensure fairness of the algorithm, we tested DICE within each demographic patient subgroups in the HF dataset. The AUC for Unknown, Asian, Other, Black, and White are 0.9053, 0.8824, 0.8563, 0.8321, 0.8470, respectively, when cluster membership is used as the predictor. The AUC for Unknown, Asian, Other, Black, and White are 0.8632, 0.8289, 0.7816, 0.8535, 0.8525, respectively, when learned representation is used as the predictor.

#### Ablation study

We conducted an ablation experiment on the HF dataset to gauge the effect of the statistical significance constraint. When we disabled the statistical significance constraint, 2 clusters were chosen based on AUC in NAS. The distribution of outcome is 80.1% and 9.01% within the two clusters, compared to the 4-level separation in Figure 3(a). The maximum AUC score is 0.8427 in the ablation study compared to the maximum AUC score 0.8539 with the statistical significance constraint. In addition, the number of neural network which met the significance constraint significantly drops from 82.4% to 64.7% for *K* = 5. These three phenomenons indicate that statistical significance constraint contributes to clearer outcome stratification especially for bigger *K*.

## 5 Discussion

An important distinction between DICE and purely unsupervised, or supervised, tasks is that DICE learns outcome-driven clusters in an un-labeled population, where the outcome-driven clusters can later be used to assign risk-levels for future unseen cohort. When there is a lack of precise treatment protocol that applies to individual patients, such as when management plans for COVID-19 were still being developed, the capability of classifying a new patient according to the learned risk-level in the seen population can provide data-driven insights in guiding subsequent management.

In this paper, We demonstrated DICE using AE for representation learning, followed by clustering of the representation using K-means, and an alternative grid search for NAS, to discover subgroups of patients in two disease populations: HF and COVID-19. We discovered that compared to baseline, DICE largely better separated the population as measured by evaluation indices for clustering. The learned representation from DICE lead to higher AUC in classifying individual outcomes, and was further used to assign unseen data into risk-levels. This technique may be used for early identification of practice and patient patterns that suggest risks, patients who may benefit from specialized care, and patients who are on trajectory for quick recovery and early discharge pathways as a form of clinical decision support.

## Broader Impact

DICE was proposed to join concepts of deep learning and statistics in healthcare to promote better acceptance of deep learning results. One of the biggest challenges to the successful application of machine learning, and especially deep learning, algorithms in healthcare is its acceptance by clinicians as an interpretable models. Traditionally, biostatistical models and concepts such as statistical significance have been better understood and accepted by clinicians and continue to be so. Thus, one implication of this work is to bridge the gap between deep learning and statistics in the context of healthcare by driving the unsupervised tasks towards statistically significant results. Aside from heart failure and COVID-19, an additional example is sepsis, which afflicts nearly million Americans each year with a mortality rate of 270,000 patients per year. The current sepsis treatment guidelines have provided a standardized approach, including aggressive intravenous hydration and early administration of antibiotics. Though significant research has been performed with resulting improvements in overall patient morbidity and mortality, much dispute still exists regarding the indiscriminate application of this protocol to all sepsis-suspected patients. In 2018, the Infectious Diseases Society of America released a statement of non-endorsement as they felt these guidelines generated more harm, particularly for those with less severe disease. Furthermore, recent evidence suggests that multiple subtypes of sepsis may exist, suggesting that this “one-size-fits-all” solution may need to reconsidered. For clinical needs such as sepsis and beyond, DICE may provide opportunities for discovery in the nuances in diagnostics and therapy while ensuring a targeted outcome.

## Data Availability

The data is internal data of Weill Cornell Medicine, and has been approved by the board of IRB in Weill Cornell Medicine.

## Notes

### Competing Interest Statement

The authors have declared no competing interest.

### Author Declarations

The study has been approved by the board of IRB in Weill Cornell Medicine.

### Summary of Updates

Adding co-authors

